# Effectiveness of the Common Elements Treatment Approach (CETA) for mental and behavioral health outcomes among women struggling to remain adherent to HIV treatment and who have experienced intimate partner violence in South Africa: A randomised controlled trial

**DOI:** 10.1101/2025.10.14.25337970

**Authors:** Amy Zheng, Jeremy C. Kane, Sithabile Mngadi-Ncube, Matthew P. Fox, Pertunia Manganye, Lawrence Long, Kristina Metz, Srishti Sardana, Michelle Alto, Ross Greener, Donald M. Thea, Laura K. Murray, Sophie Pascoe

## Abstract

**Background:** Rates of intimate partner violence (IPV) and HIV in South Africa are among the highest globally. IPV is associated with a range of adverse mental health and HIV outcomes. The Common Elements Treatment Approach (CETA) is a transdiagnostic, evidence-based intervention delivered by lay providers.

**Objective:** To compare the effectiveness of CETA to active attention control in reducing IPV, depression, Post-Traumatic Stress Disorder (PTSD), and substance use among women at risk of poor HIV outcomes who have experienced IPV.

**Methods:** Women living with HIV with an unsuppressed viral load or at risk for poor adherence and experienced past 12-month IPV were recruited from Johannesburg-area clinics and randomised 1:1 to CETA or control (SMS HIV appointment reminders plus safety checks and planning). The primary trial outcome was HIV retention and viral suppression, reported elsewhere. This paper reports secondary outcomes, evaluated at three and 12 months: IPV, depression, PTSD, and substance use.

**Findings:** Participants were enrolled between November 11, 2021 to July 19, 2023 and randomised to CETA (N=202) or control (N=197). In the intent to treat analysis, the Cohen’s d treatment effect for depression at three months was 0.24 (difference in mean change -3.1; 95% CI: -6.1, 0.1) and 0.48 at 12 months (-6.2; 95% CI: -9.5, -2.8). The PTSD treatment effect was 0.39 at three (-0.3; 95% CI: -0.5, -0.1) and 0.47 at 12 months (-0.3; 95% CI: -0.5, -0.2). Effect sizes were larger in a subgroup of participants with the top 50% of baseline symptom scores (depression: d=0.50, d=0.74; PTSD: d=0.58, d=0.94, at three and 12 months, respectively). There were no statistically significant differences in change for substance use or IPV. At baseline, only 12% of participants had past 3-month substance use and 32% had past 3-month or ongoing experiences of IPV, which made these outcomes challenging to evaluate.

**Conclusions:** CETA was effective for reducing depression and PTSD including among high severity participants and at an extended follow-up. Future studies with increased power for substance use and IPV outcomes are warranted.

**Clinical implications:** CETA is a recommended treatment for depression and PTSD among this population.

**Trial registration number:** Clinicaltrials.gov NCT04242992, registered January 27, 2020

**Key Messages:** *What is already known about this topic?:* Intimate partner violence (IPV) and related mental health problems are common in South Africa and can lead to poor HIV outcomes, such as low retention in care and viral non-suppression. There is a lack of evidence-based mental healthcare options for women living with HIV who have experienced IPV.

*What this study adds:* Among women living with HIV and past-year IPV experiences, we found that Common Elements Treatment Approach (CETA) was an effective treatment for depression and PTSD compared to a control condition.

*How this study might affect research, practice, or policy:* CETA is recommended to treat common mental health problems among women with HIV and experiences of IPV.

## INTRODUCTION

In South Africa, almost eight million people are living with HIV (PWH) [1]. To achieve epidemic control, South Africa aims to achieve the UNAIDS 95-95-95 targets by 2030 whereby 95% of PWH know their HIV status, 95% of people who know their status are in care and receiving antiretroviral therapy (ART), and 95% of people on ART are virally suppressed [2]. As of 2024, South Africa has achieved the first target with 95% of the eight million PWH aware of their status; however, progress towards the other two targets have slowed with 81% of those who know their status receiving ART and 92% of those on ART virally suppressed [3].

Certain populations remain well below the 95-95-95 targets, including women who have experienced intimate partner violence (IPV). A recent pooled analysis of 57 surveys in sub-Saharan Africa found that women who experienced IPV in the past year were less likely to be virally suppressed than those who had not experienced IPV [4]. This is a particular concern in South Africa, where up to 50% of women experience IPV in their lifetime [5]. Experiences of IPV are associated with increased risk for mental and behavioral health problems, including depression, post-traumatic stress disorder (PTSD), and substance use [6], which are themselves associated with poor HIV outcomes [7]. Addressing mental health and IPV is critical to achieving the 95-95-95 targets in South Africa. However, there is a substantial mental health treatment gap in South Africa similar to most low-and-middle-income countries (LMIC): between 75% and 92% of individuals in need of mental health care do not receive treatment and the quality of care that does exist is variable [8].

The Common Elements Treatment Approach (CETA) is an evidence-based, transdiagnostic intervention comprised of cognitive-behavioral therapy (CBT) components [9]. CETA was designed to address a variety of commonly co-occurring mental and behavioral health problems including depression, PTSD, violence, and substance use. CETA can be delivered by lay counselors at lower cost than traditional psychological treatments thereby addressing a critical implementation barrier in LMIC. In diverse LMIC contexts and populations, CETA has been found effective at treating a range of mental health problems in randomized controlled trials (RCTs) [10–14]. In a trial of women experiencing IPV perpetrated by a current male partner with unhealthy alcohol use, CETA reduced IPV substantially when both women and men received CETA [15]. CETA has the potential to improve HIV treatment outcomes by addressing underlying barriers to retention and adherence (e.g., IPV and other co-occurring mental health conditions). However, CETA has not been tested for mental health outcomes within the context of HIV and IPV in South Africa, and prior trials were also limited by short (i.e., ≤ 6 months) follow-up.

## OBJECTIVE

We conducted a RCT of CETA among women in Johannesburg, South Africa who were at-risk of poor retention and/or viral non-suppression and reported experiencing IPV in the past year. The primary outcome of the trial was a combined retention in care/viral suppression, and those results will be disseminated in a forthcoming publication. This paper reports on the effectiveness of CETA at reducing mental and behavioral health problems (depression, PTSD, substance use) and IPV, which were secondary trial outcomes, compared to an active control condition.

## METHODS

### Setting

This was an individual, parallel RCT testing the effectiveness of CETA compared to an active attention control condition. The study was conducted at two large public sector HIV care and treatment sites in Johannesburg, South Africa. Women were recruited from the clinic after initially being referred by clinic staff who believed they might meet the inclusion criteria. Study staff provided a brief overview of the study and then conducted an initial abbreviated consent process for screening to confirm the inclusion/exclusion criteria. The abbreviated consent process explained questions participants would be asked, including questions on IPV.

### Participants

Women were eligible for the trial if they met the following criteria:

- 18 years old
- Living with HIV and on ART
- Had at least one of the following:
  - Unsuppressed viral load (>50 copies/mL)
  - Defaulted from treatment since their last viral load

- Missed or late (>14 days) clinic appointment in the past year
- Had *any* past 12-month experience of IPV as reported on the Severity of Violence Against Women Scale (SVAWS) [17]
- Were literate and able to speak and read English, Zulu or SeSotho.

We excluded women who: 1) were currently psychotic or on an unstable psychiatric regimen; 2) had a suicide attempt/ideation with intent and plan and/or self-harm in the past month; or 3) were enrolled in any other HIV treatment intervention study.

The IPV inclusion criterion was evaluated using a self-administered tablet-based version of the SVAWS physical/sexual violence subscale. Eligibility was defined as *any* IPV occurring in the past 12 months at the time of assessment. The SVAWS physical/sexual violence subscale consists of 27-items asking about the frequency of violence perpetuated by their male partner. All women who were screened were given resources for hotlines and local programs supporting women experiencing IPV. Women who screened as eligible (e.g., consented to the abbreviated process and indicated any past 12-month IPV) then completed a full consent process that provided a detailed overview of the study including: what the intervention and control arms were, their chances of receiving the intervention, follow-up procedures, safety monitoring, risks and benefits of the study, and permission to allow the study team access to their medical records for HIV-treatment outcomes at 12- and 24-months. Those who enrolled in the study completed a baseline assessment using an audio computer-assisted self-interview (ACASI) tablet-based questionnaire in any of the three languages (see Measurement and outcomes) and then randomised (see Randomisation).

### Interventions

#### CETA

Participants randomised to the intervention arm received CETA, a modular, transdiagnostic, flexible therapy approach implemented by lay counselors. CETA is comprised of nine evidence-based CBT elements to treat a variety of common problems, including violence, substance use, depression, anxiety, trauma-related symptoms and other behaviors that increase vulnerability (e.g., unprotected sex, medication non-adherence). CETA’s multi-problem, flexible approach addresses comorbidities and allows for individualized treatment [9].

Twelve lay counsellors were trained via a ten day in-person training, and then participated in supervision groups, led by a local supervisor. Groups of 5-6 counsellors met weekly with a supervisor for two hours to review cases, present problems, and decide which treatment elements should be used for each participant. Each supervisor had a weekly call with a CETA trainer to ensure treatment fidelity, provide close oversight of high-risk safety cases, and address any implementation difficulties.

Those randomized to CETA were assigned a counsellor who provided 6-12 CETA sessions, aiming for these to be done on a weekly basis for 60 minutes each. Women were asked if they wanted to invite a male partner to participate in CETA. Only 15 male partners were invited and participated in CETA with a separate counsellor. Sessions included Encouraging Participation/Introduction (i.e. learning about the symptoms and program), Cognitive Coping/Restructuring, Gradual Exposure for Trauma, Behavioral Activation, Substance Use Reduction, Problem Solving, and Relaxation and Safety. The elements implemented, dosing, and number of sessions were dependent on the client’s clinical presentation. Due to COVID-19, which was widespread in South Africa during this trial, as well as challenges women reported in getting to the clinic (i.e., distance and time to the clinic), we allowed for CETA to be delivered telephonically, although few participants chose this option (n = 23; 11.5%).

At each session participants were asked about their safety related to any ongoing IPV. If a session was not held for that week, participants were contacted via SMS or phone call. Participants were reimbursed ZAR50 (∼USD3) for transportation costs incurred to attend CETA sessions.

#### Control

The active control condition included weekly SMS to participants that provided reminders about their HIV care appointments and asked questions on their safety (i.e., suicidal and homicidal intent and IPV) over the past week. These SMS questions were sent for 12 weeks. In addition to weekly SMS messages, participants received usual care referrals for any mental health problems they were experiencing.

#### Safety for both arms

Given the potential high-risk nature of a study population experiencing IPV in the past 12 months, participants in both arms received safety monitoring, including assessment and planning. At enrollment, counsellors met with each woman to assess their IPV risk and develop a safety plan as needed. Counsellors checked in weekly with all participants for safety assessments and asked: 1) Are you thinking of killing yourself? 2) Do you have a plan to kill yourself? 3) Do you have the means to kill yourself? and 4) Are you at risk of serious injury or death from interpersonal violence? For control participants, this was completed via SMS. For CETA participants, this was completed during the weekly sessions (either in-person or via the telephone). If a session was missed, participants were telephoned to complete the safety questions and additional planning, as needed. Additional precautions were taken to ensure client safety when completing these questions via SMS or phone, including ensuring privacy during telephone contact and/or personalizing and coding SMS messages. If a participant responded yes to any of the questions, further safety assessment and immediate intensive safety planning was completed either via phone or in-person by a trained CETA counsellor. CETA trainers were contacted within 24 hours for all safety cases to ensure that local safety protocols were followed.

### Measurement and outcomes

Mental health and IPV outcomes were evaluated actively through ACASI, which allowed participants to read questions on a laptop screen and simultaneously hear the questions being read to them while wearing headphones. Text and audio were available in the language of the participant’s choice. In prior research, ACASI has reduced social desirability bias [18], which we believed was important given the sensitive nature of the outcomes. The ACASI-based assessment was administered at baseline, and three- and 12-months post-baseline. Participants received ZAR150 (∼USD8) upon completion of each assessment. Because our primary outcome was retention based, we made no attempt to retain patients in the study nor ensure they attended a follow-up assessment (except for safety support which continued even for participants who missed visits) beyond standard outreach to complete the CETA intervention and follow-up assessments as any effort to do so would undermine the primary outcome (e.g., whether CETA improves retention and viral suppression).

The outcomes assessment included:

- Severity of Violence Against Women Scale (SVAWS) [17], a 46-item questionnaire evaluating the frequency and severity of IPV. The scale consisted of three subscales: threatened violence (19 items), physical violence (21 items), and sexual violence (6 items). Like the prior trial of CETA’s effectiveness on IPV in Zambia [15], we analyzed the threatened violence subscale and a combined physical/sexual violence subscale as two discreet outcomes. At the three-month and 12-month follow-up assessments, we modified the reference period for the scale to be the past three months (from past 12-months at baseline). Continuous subscale scores with higher scores indicating greater severity IPV were used as outcomes (threatened scale possible range: 19-76; physical/sexual IPV scale possible range: 27-108).
- Center for Epidemiological Studies-Depression Scale (CES-D) [19], a 20-item scale of past week depression symptoms. A continuous depression score (possible range 0-60) with higher score indicating greater severity depression was used as the outcome.
- Harvard Trauma Questionnaire (HTQ) [20], a 39-item scale of past week PTSD symptoms. A continuous PTSD score (possible range 1-4) with higher score indicating greater severity PTSD was used as the outcome.
- Alcohol, Smoking, and Substance Involvement Screening Test (ASSIST) [21] which evaluates past three-month substance use for a range of substance types. We derived a binary variable of any past three-month use as the outcome for analysis.

### Randomisation

After the baseline assessment, participants were immediately randomised 1:1 to CETA or control using blinded blocked randomisation through a computer accessed, randomisation code (generated by the US-based investigative team) accessed via a tablet by study staff. Participants were immediately notified of the randomisation result. As the intervention required specific counselling, neither counsellors nor participants were blinded to their study arm, but data capture and analysis was blinded with unblinding only after data analysis was completed on three- and 12-month month mental health outcomes (and after all data, including HIV data, were collected for the primary 12-month outcome).

### Sample Size

Based on findings from prior studies [22] and assuming 80% power with a 2-sided α= 0.05, a sample of 182 total (91/treatment arm) was required to detect a 20-percentage point increase in the proportion suppressed and retained between the CETA and control arms. The sample size was then increased to 400 total (200/treatment arm) to allow for sufficient power to evaluate mediators and moderators. Based on power calculations from previous CETA trials [15,23], we also believed this sample size was sufficient to estimate at least a medium effect size (Cohen’s d=0.5) for the mental health and IPV outcomes.

### Statistical Analysis

Primary analyses were by intent-to-treat (ITT) and included all enrolled participants who were not disenrolled after randomisation (Supplemental Figure 1). We used multiple imputation (20 sets of imputations) with chained equations to assign values to missing data. We used linear mixed effects regression models to assess the effect of CETA compared to the control group for continuous outcomes: IPV (threatened and physical/sexual), depression, and PTSD. We used a generalized linear mixed effect model for the binary outcome of any substance use. Treatment group, time, and an interaction term between treatment group and time were included as fixed effects with a random effect for participant ID to account for repeat measures of the same participant over time. Robust standard errors were estimated for each outcome using the *vce(robust)* option in Stata [24]. The interaction term in the linear models represents the mean difference in change from baseline to each follow-up assessment between CETA and control. For all continuous outcomes, we calculated Cohen’s *d* effect sizes by dividing the effect of the intervention (as indicated by the beta coefficient for the interaction term) by the pooled baseline standard deviation. An effect size ranging from 0.2-0.4 indicated a small effect, >0.4-0.7 a medium sized effect, and >0.7 a large effect [25]. The interaction term in the generalized linear model represented a change in risk between CETA and control from baseline to each follow-up. All analyses were conducted in Stata v16 [24].

In addition to the ITT analysis, we conducted the following secondary analyses: 1) a per-protocol analysis where we excluded participants who did not complete CETA, and 2) a subgroup analysis of participants in the top 50% of baseline symptoms for the IPV, depression, and PTSD outcomes regardless of treatment arm to explore the effectiveness of CETA among participants with the highest severity problems.

### Ethics Approval

The trial was prospectively registered on Clincialtrials.gov (NCT04242992) on January 24, 2020. The trial protocol was previously published [16]. The study protocol was approved by the University of the Witwatersrand Human Research Ethics Committee (Medical) (M200353), the Boston University Institutional Review Board (H-39746), the Johns Hopkins School of Public Health Institutional Review Board (12546), and the Columbia University Institutional Review Board (AAAS9661). Written informed consent was obtained from all study participants prior to enrolment by study staff. The trial was monitored by a 3-person Data and Safety Monitoring Board (DSMB). The results are reported according to CONSORT guidelines (see supplement for CONSORT checklist).

## RESULTS

Study enrollment took place from November 11, 2021 to July 19, 2023. Two hundred and two participants were randomized to the CETA Arm and 197 were randomized to the control arm (Supplemental Figure 1). Of the 202 randomized to the CETA Arm, 144 (71.3%) completed CETA and averaged 7.6 sessions (SD 5.5) over an average of 12 (SD 7.0) weeks (Supplemental Figure 1). The overall mean age was 40.6 years (SD: 8.8). Most participants were of South African nationality, unemployed but looking for work, reported never being married, and experiencing high levels of food insecurity (Table 1). Although all participants reported experiencing IPV in the past year (as an inclusion criterion), only approximately one-third reported past 3-month or ongoing IPV (31.5%; n=126) and 36.6% (n=146) were living with the IPV perpetrator. Baseline characteristics were similar when stratified by treatment arm.

**Table 1.** Baseline demographics of individuals enrolled in a randomized trial of the effect of CETA on mental health outcomes in Johannesburg South Africa among women with HIV on ART who have experienced violence and challenges with adherence by treatment arm (N = 399)^1,2^.

|  | **CETA Arm**<br>**(n = 202)** | **Control Arm**<br>**(n = 197)** | **Total**<br>**(n = 399)** |
| --- | --- | --- | --- |
| **Age, mean (SD)** | 40.4 (8.3) | 40.9 (9.3) | 40.6 (8.8) |
| **Race** |  |  |  |
| *Black African* | 196 (97.0) | 192 (97.5) | 388 (97.2) |
| *Coloured* | 2 (1.0) | 5 (2.5) | 7 (1.8) |
| *Missing* | 4 (2.0) | 0 | 4 (1.0) |
| **Education** |  |  |  |
| *Grade 7 or less* | 12 (6.0) | 7 (3.6) | 19 (4.8) |
| *Grade 8-10* | 30 (14.9) | 30 (15.2) | 60 (15.0) |
| *Grade 11-12* | 105 (52.0) | 101 (51.3) | 206 (51.6) |
| *Completed Grade School* | 34 (16.8) | 45 (22.8) | 79 (19.8) |
| *Certificate/Diploma/Post-Secondary* | 12 (5.9) | 13 (6.6) | 25 (6.3) |
| *Degree* | 4 (2.0) | 1 (0.5) | 5 (1.3) |
| *Unknown/Missing* | 5 (2.5) | 0 | 5 (1.3) |
| **Currently in School** | 14 (6.9) | 17 (8.6) | 31 (7.8) |
| **Employment** |  |  |  |
| *Unemployed and Looking for Work* | 141 (69.8) | 120 (60.9) | 261 (65.4) |
| *Unemployed and Not Looking for Work* | 5 (2.5) | 3 (1.5) | 8 (2.0) |
| *Self-Employed (Part- or Full-Time)* | 12 (6.0) | 21 (10.7) | 33 (8.3) |
| *Employed Part-Time* | 23 (11.4) | 29 (14.7) | 52 (13.0) |
| *Employed Full-Time* | 15 (7.4) | 21 (10.7) | 36 (9.0) |
| *Other/Missing* | 6 (3.0) | 3 (1.5) | 9 (2.3) |
| **Currently living with perpetrator of IPV** | 75 (37.1) | 71 (36.0) | 146 (36.6) |
| **Marital Status** |  |  |  |
| *Never Married* | 147 (72.8) | 142 (72.1) | 289 (72.4) |
| *Married* | 35 (17.3) | 30 (15.2) | 65 (16.3) |
| *Divorced/Separated* | 13 (6.5) | 14 (7.2) | 27 (6.8) |
| *Widowed* | 4 (2.0) | 11 (5.6) | 15 (3.8) |
| *Unknown/Missing* | 3 (1.5) | 0 | 3 (0.8) |
| **Have a Current Partner** | 178 (88.1) | 181 (91.9) | 359 (90.0) |
| **Have Children** | 176 (87.1) | 172 (87.3) | 348 (87.7) |
| **Food Insecurity** |  |  |  |
| *Never* | 66 (32.7) | 47 (23.9) | 113 (28.3) |
| *Seldom* | 13 (6.4) | 20 (10.2) | 33 (8.3) |
| *Sometimes* | 105 (52.0) | 127 (64.5) | 232 (58.2) |
| *Often* | 12 (5.9) | 3 (1.5) | 15 (3.8) |
| *Missing* | 6 (3.0) | 0 | 6 (1.5) |
| **Financial Status** |  |  |  |
| *Very Difficult* | 74 (36.6) | 63 (32.0) | 137 (34.3) |
| *Difficult* | 96 (47.5) | 106 (53.8) | 202 (50.6) |
| *Easy* | 26 (12.9) | 26 (13.2) | 52 (13.0) |
| *Very Easy* | 1 (0.5) | 2 (1.0) | 3 (0.8) |
| *Missing* | 5 (2.5) | 0 | 5 (1.3) |
| **Suicidal Ideation** | 92 (45.5) | 73 (37.1) | 165 (41.4) |
| **Homicidal Ideation** | 47 (23.3) | 43 (21.9) | 90 (22.3) |
| **SVAWS Threatened Score, mean (SD) [range]** | 38.9 (13.7) [20-73] | 37.6 (12.4) [19-68] | 38.2 (13.1) [19-73] |
| **SVAWS Physical Score, mean (SD) [range]** | 61.6 (23.2) [34-125] | 59.6 (20.3) [34-107] | 60.6 (21.8) [34-125] |
| **CES-D Score, mean (SD) [range]** | 31.8 (13.0) [5-53] | 29.2 (12.7) [3-54] | 30.5 (12.9) [21-45] |
| **HTQ Score, mean (SD) [range]** | 2.7 (0.7) [2.1-3.3] | 2.5 (0.7) [2.0-3.1] | 2.6 (0.7) [2.1-3.2] |
| **Any past three-month illicit substance use** | 28 (13.9) | 19 (9.6) | 47 (11.8) |
| **Missed a Visit within the Past Year<sup>3</sup>** | 122 (60.4) | 119 (60.4) | 241 (60.4) |
| **Late to a Visit within the Past Year<sup>3</sup>** | 69 (34.2) | 54 (27.4) | 123 (30.8) |
| **Defaulted on Treatment within the Past Year<sup>3</sup>** | 114 (56.4) | 113 (57.4) | 227 (56.9) |
| **Years on ART, mean (SD)** | 9.0 (5.7) | 8.5 (5.7) | 8.7 (5.7) |
| **Viral Load, copies/mL, median (IQR)** | **30 (20, 107.5)** | **39 (20, 139.5)** | **35 (20, 110)** |
| *0 – 50* | 125 (61.9) | 122 (61.9) | 247 (61.9) |
| *51 - 500* | 44 (21.8) | 38 (19.3) | 82 (20.6) |
| *501 - 1,000* | 5 (2.5) | 3 (1.5) | 8 (2.0) |
| *1,001 - 5,000* | 8 (4.0) | 7 (3.6) | 15 (3.8) |
| *5,001 - 50,000* | 6 (3.0) | 8 (4.1) | 14 (3.5) |
| *50,001 - 500,000* | 2 (1.0) | 9 (4.6) | 11 (2.8) |
| *500,001+* | 2 (1.0) | 1 (0.5) | 3 (0.8) |
| *Missing* | 10 (5.0) | 9 (4.6) | 19 (4.8) |

CETA= Common Elements Treatment Approach; Control Arm received 12 weeks of weekly SMS messaging; SVAWS = Severity of Violence Against Women Scale, CES-D = Center for Epidemiologic-Studies Depression Scale; HTQ = Harvard Trauma Questionnaire.
<sup>1</sup>Based off the non-imputed dataset
<sup>2</sup>Total N = 399 instead of 400 because there were individuals who were randomized and then disenrolled after randomisation. See Figure 1 for more details.
<sup>3</sup>May sum to greater than the total sample size across these three markers of engagement with care as these are non-mutually exclusive categories

Table 2 presents the intent-to-treat intervention effect for IPV, depression, PTSD and substance use. The Cohen’s d treatment effect of threatened IPV was 0.15 at three-months (difference in Mean Change: -2.0; 95% Confidence Interval (CI): -4.6, 0.7) and 0.18 at 12-months (-2.4; 95% CI: -5.2, 0.4) suggesting almost no effect of CETA on threatened IPV. For physical and sexual IPV, we found a treatment effect of 0.08 at three-months (-1.7; 95% CI: -6.2, 2.8) and 0.18 at 12-months (-3.9; 95% CI: -8.5, 0.7), again suggesting almost no effect at three-months and a potentially small effect by CETA on physical and sexual IPV at 12-months. However, 38.2% (n = 71) had not yet completed treatment at the time of the 3-month assessment and 8.6% (n = 16) completed treatment on the same day as the 3-month assessment. Similarly, 27.4% (n = 51) had not yet completed treatment at the time of the 12-month assessment and 1.1% (n = 2) completed treatment on the same day as the 12-month assessment

**Table 2.**
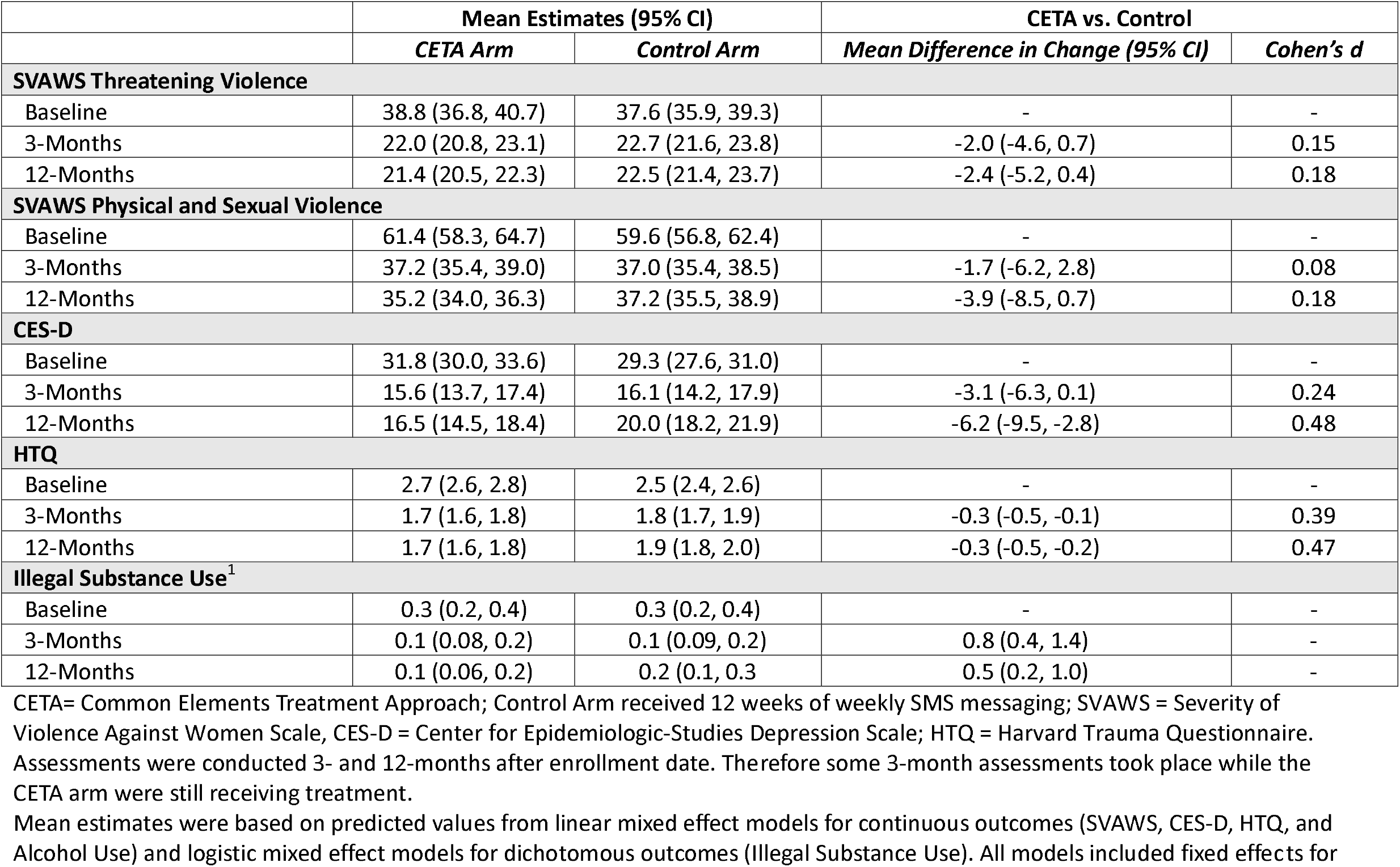
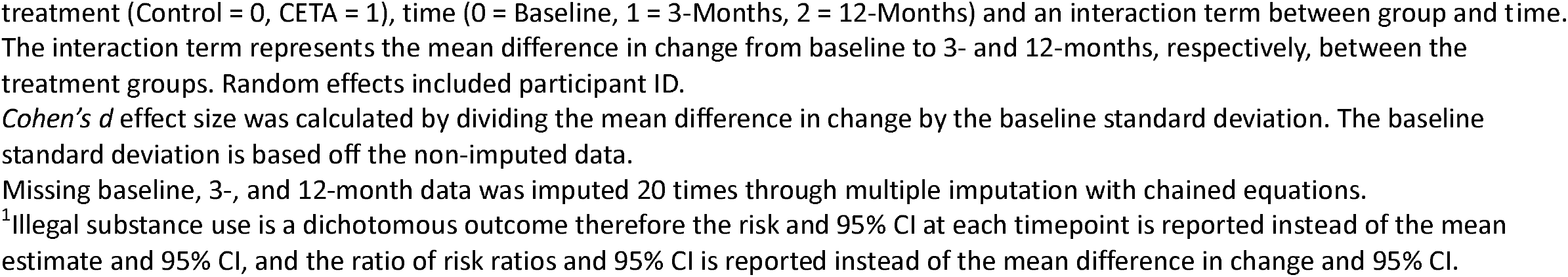
Estimated mean outcome differences and clinical effect sizes among the full study sample in a randomized trial of the effect of CETA on mental health outcomes in Johannesburg South Africa among women with HIV on ART who have experienced violence and challenges with adherence (N = 399)

|  | Mean Estimates (95% CI) |  | CETA vs. Control |  |
| --- | --- | --- | --- | --- |
|  | CETA Arm | Control Arm | Mean Difference in Change (95% CI) | Cohen's d |
| **SVAWS Threatening Violence** |  |  |  |  |
| Baseline | 38.8 (36.8, 40.7) | 37.6 (35.9, 39.3) | - | - |
| 3-Months | 22.0 (20.8, 23.1) | 22.7 (21.6, 23.8) | -2.0 (-4.6, 0.7) | 0.15 |
| 12-Months | 21.4 (20.5, 22.3) | 22.5 (21.4, 23.7) | -2.4 (-5.2, 0.4) | 0.18 |
| **SVAWS Physical and Sexual Violence** |  |  |  |  |
| Baseline | 61.4 (58.3, 64.7) | 59.6 (56.8, 62.4) | - | - |
| 3-Months | 37.2 (35.4, 39.0) | 37.0 (35.4, 38.5) | -1.7 (-6.2, 2.8) | 0.08 |
| 12-Months | 35.2 (34.0, 36.3) | 37.2 (35.5, 38.9) | -3.9 (-8.5, 0.7) | 0.18 |
| **CES-D** |  |  |  |  |
| Baseline | 31.8 (30.0, 33.6) | 29.3 (27.6, 31.0) | - | - |
| 3-Months | 15.6 (13.7, 17.4) | 16.1 (14.2, 17.9) | -3.1 (-6.3, 0.1) | 0.24 |
| 12-Months | 16.5 (14.5, 18.4) | 20.0 (18.2, 21.9) | -6.2 (-9.5, -2.8) | 0.48 |
| **HTQ** |  |  |  |  |
| Baseline | 2.7 (2.6, 2.8) | 2.5 (2.4, 2.6) | - | - |
| 3-Months | 1.7 (1.6, 1.8) | 1.8 (1.7, 1.9) | -0.3 (-0.5, -0.1) | 0.39 |
| 12-Months | 1.7 (1.6, 1.8) | 1.9 (1.8, 2.0) | -0.3 (-0.5, -0.2) | 0.47 |
| **Illegal Substance Use<sup>1</sup>** |  |  |  |  |
| Baseline | 0.3 (0.2, 0.4) | 0.3 (0.2, 0.4) | - | - |
| 3-Months | 0.1 (0.08, 0.2) | 0.1 (0.09, 0.2) | 0.8 (0.4, 1.4) | - |
| 12-Months | 0.1 (0.06, 0.2) | 0.2 (0.1, 0.3) | 0.5 (0.2, 1.0) | - |

CETA= Common Elements Treatment Approach; Control Arm received 12 weeks of weekly SMS messaging; SVAWS = Severity of Violence Against Women Scale, CES-D = Center for Epidemiologic-Studies Depression Scale; HTQ = Harvard Trauma Questionnaire. Assessments were conducted 3- and 12-months after enrollment date. Therefore some 3-month assessments took place while the CETA arm were still receiving treatment.
Mean estimates were based on predicted values from linear mixed effect models for continuous outcomes (SVAWS, CES-D, HTQ, and Alcohol Use) and logistic mixed effect models for dichotomous outcomes (Illegal Substance Use). All models included fixed effects for
treatment (Control = 0, CETA = 1), time (0 = Baseline, 1 = 3-Months, 2 = 12-Months) and an interaction term between group and time. The interaction term represents the mean difference in change from baseline to 3- and 12-months, respectively, between the treatment groups. Random effects included participant ID.
*Cohen's d* effect size was calculated by dividing the mean difference in change by the baseline standard deviation. The baseline standard deviation is based off the non-imputed data.
Missing baseline, 3-, and 12-month data was imputed 20 times through multiple imputation with chained equations.
<sup>1</sup>Illegal substance use is a dichotomous outcome therefore the risk and 95% CI at each timepoint is reported instead of the mean estimate and 95% CI, and the ratio of risk ratios and 95% CI is reported instead of the mean difference in change and 95% CI.

The treatment effect for depression was small at 0.24 at three-months (-3.1; 95% CI: -6.1, 0.1). By 12 months, the treatment effect increased to 0.48 (-6.2; 95% CI: -9.5, -2.8) showing not just a sustained but a larger effect on depression. For PTSD the treatment effect was 0.39 at three months (-0.3; 95% CI: -0.5, -0.1), and 0.47 at 12-months (-0.3; 95% CI: -0.5, -0.2) again showing a sustained, medium effect size by CETA on PTSD. For substance use the ratio of risk ratios was 0.8 at three-months (95% CI: 0.4, 1.4) and was 0.5 at 12-months (95% CI: 0.2, 1.0). This suggests that substance use reduced to a greater degree among participants who received CETA compared to control, however, these findings were not statistically significant.

Table 3 presents the per protocol analysis that excluded participants who did not complete CETA (n=56). In this sub-sample, the treatment effect of threatened IPV was small at 0.26 at three-months (-3.3; 95% CI: -6.2, -0.4) and 0.23 at 12-months (-2.9; 95% CI: -6.0, 0.1) unlike the intent-to-treat results. Similarly, for physical and sexual IPV, we found an imprecise, but small treatment effect at three (Cohen’s *d*: 0.15) and 12 (Cohen’s *d*: 0.19) months. Treatment effects for depression were greater when compared to the intent-to-treat results at both time points with an effect of 0.41 at three-months (-5.3; 95% CI: -8.7, -1.9) and 0.52 at 12-months (-6.7; 95% CI: -10.6, -2.8). For PTSD we saw a sustained, medium sized effect of the intervention at 0.53 at three-months (-0.4; 95% CI: -0.6, -0.2) and 0.54 at 12-months (-0.4; 95% CI: -0.6, -0.2). Lastly, for the outcome of any substance use the at three-months the ratio of risk ratios was 0.8 (95% CI: 0.5, 1.2) and at 12-months the ratio of risk ratios was 0.6 (95% CI: 0.4, 0.9).

**Table 3.**
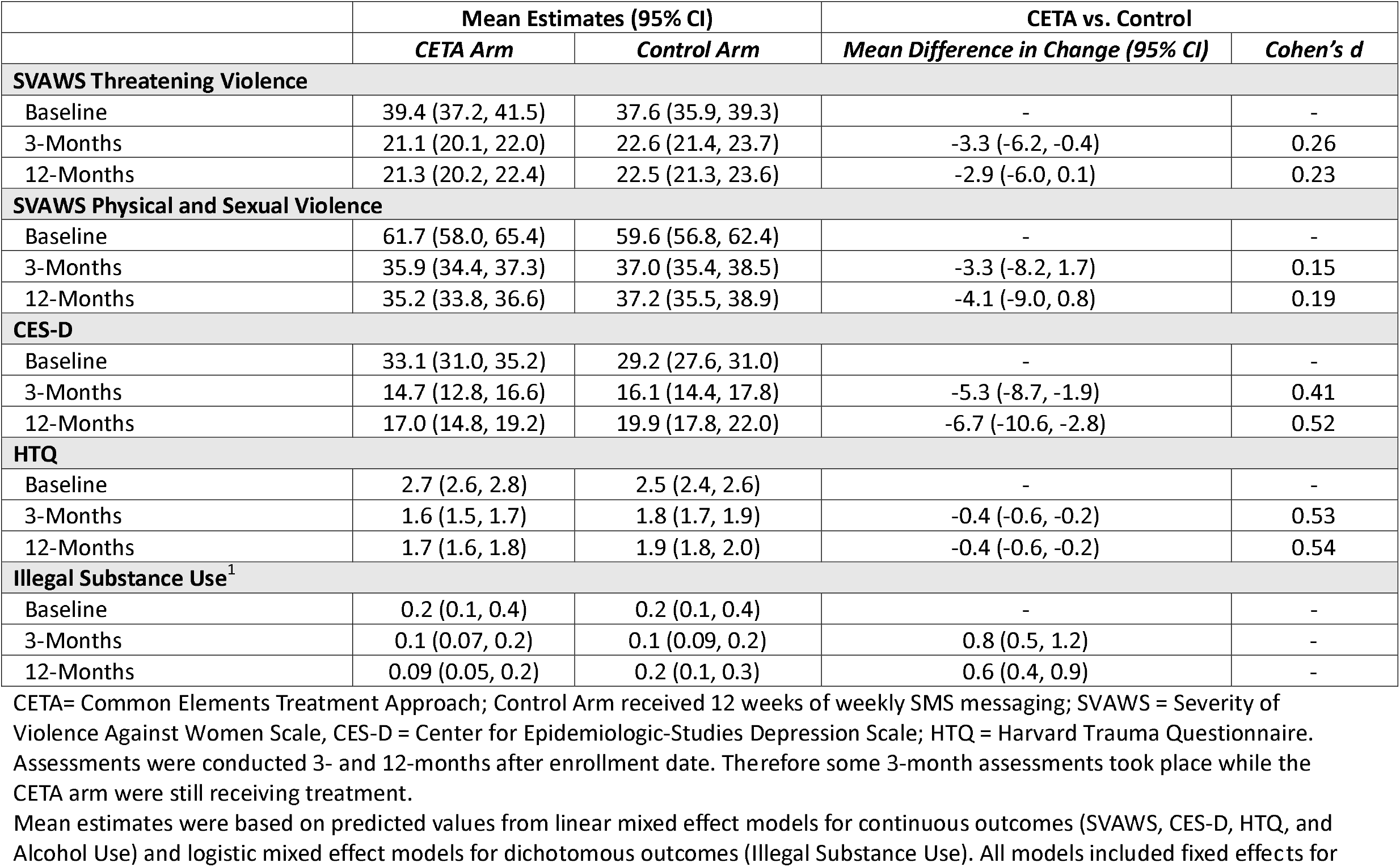
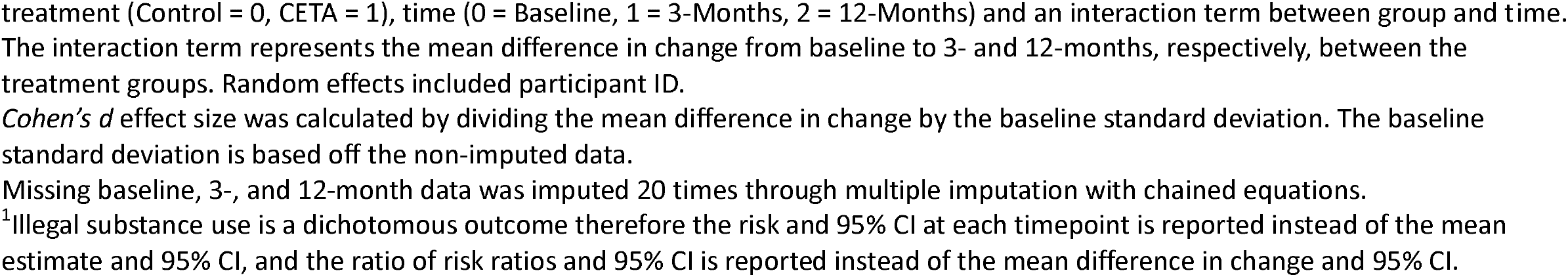
Per-Protocol Analysis 1: Estimated mean outcome differences and clinical effect sizes among participants who completed CETA in a randomized trial of the effect of CETA on mental health outcomes in Johannesburg South Africa among women with HIV on ART who have experienced violence and challenges with adherence (N = 343)

|  | Mean Estimates (95% CI) |  | CETA vs. Control |  |
| --- | --- | --- | --- | --- |
|  | CETA Arm | Control Arm | Mean Difference in Change (95% CI) | Cohen's d |
| **SVAWS Threatening Violence** |  |  |  |  |
| Baseline | 39.4 (37.2, 41.5) | 37.6 (35.9, 39.3) | - | - |
| 3-Months | 21.1 (20.1, 22.0) | 22.6 (21.4, 23.7) | -3.3 (-6.2, -0.4) | 0.26 |
| 12-Months | 21.3 (20.2, 22.4) | 22.5 (21.3, 23.6) | -2.9 (-6.0, 0.1) | 0.23 |
| **SVAWS Physical and Sexual Violence** |  |  |  |  |
| Baseline | 61.7 (58.0, 65.4) | 59.6 (56.8, 62.4) | - | - |
| 3-Months | 35.9 (34.4, 37.3) | 37.0 (35.4, 38.5) | -3.3 (-8.2, 1.7) | 0.15 |
| 12-Months | 35.2 (33.8, 36.6) | 37.2 (35.5, 38.9) | -4.1 (-9.0, 0.8) | 0.19 |
| **CES-D** |  |  |  |  |
| Baseline | 33.1 (31.0, 35.2) | 29.2 (27.6, 31.0) | - | - |
| 3-Months | 14.7 (12.8, 16.6) | 16.1 (14.4, 17.8) | -5.3 (-8.7, -1.9) | 0.41 |
| 12-Months | 17.0 (14.8, 19.2) | 19.9 (17.8, 22.0) | -6.7 (-10.6, -2.8) | 0.52 |
| **HTQ** |  |  |  |  |
| Baseline | 2.7 (2.6, 2.8) | 2.5 (2.4, 2.6) | - | - |
| 3-Months | 1.6 (1.5, 1.7) | 1.8 (1.7, 1.9) | -0.4 (-0.6, -0.2) | 0.53 |
| 12-Months | 1.7 (1.6, 1.8) | 1.9 (1.8, 2.0) | -0.4 (-0.6, -0.2) | 0.54 |
| **Illegal Substance Use<sup>1</sup>** |  |  |  |  |
| Baseline | 0.2 (0.1, 0.4) | 0.2 (0.1, 0.4) | - | - |
| 3-Months | 0.1 (0.07, 0.2) | 0.1 (0.09, 0.2) | 0.8 (0.5, 1.2) | - |
| 12-Months | 0.09 (0.05, 0.2) | 0.2 (0.1, 0.3) | 0.6 (0.4, 0.9) | - |

CETA= Common Elements Treatment Approach; Control Arm received 12 weeks of weekly SMS messaging; SVAWS = Severity of Violence Against Women Scale, CES-D = Center for Epidemiologic-Studies Depression Scale; HTQ = Harvard Trauma Questionnaire. Assessments were conducted 3- and 12-months after enrollment date. Therefore some 3-month assessments took place while the CETA arm were still receiving treatment.
Mean estimates were based on predicted values from linear mixed effect models for continuous outcomes (SVAWS, CES-D, HTQ, and Alcohol Use) and logistic mixed effect models for dichotomous outcomes (Illegal Substance Use). All models included fixed effects for
treatment (Control = 0, CETA = 1), time (0 = Baseline, 1 = 3-Months, 2 = 12-Months) and an interaction term between group and time. The interaction term represents the mean difference in change from baseline to 3- and 12-months, respectively, between the treatment groups. Random effects included participant ID.
*Cohen's d* effect size was calculated by dividing the mean difference in change by the baseline standard deviation. The baseline standard deviation is based off the non-imputed data.
Missing baseline, 3-, and 12-month data was imputed 20 times through multiple imputation with chained equations.
<sup>1</sup>Illegal substance use is a dichotomous outcome therefore the risk and 95% CI at each timepoint is reported instead of the mean estimate and 95% CI, and the ratio of risk ratios and 95% CI is reported instead of the mean difference in change and 95% CI.

Table 4 presents the treatment effects after restricting analyses to participants in the top 50% of baseline symptoms for each outcome regardless of treatment arm. When compared to the intent-to-treat effect results, the effect of CETA on threatened IPV at three-months was similar when restricted to the top 50% of baseline threatened IPV scores (ITT Cohen’s *d:* 0.15; Top 50% Cohen’s *d:* 0.19). However, by 12 months results from this restricted analysis found a small treatment effect of 0.32 (-3.1; 95% CI: -6.6, 0.4) compared to the ITT’s effect of 0.18. Unlike the ITT analysis which found almost no effect for physical and sexual IPV at both timepoints, when limited to those in the top 50% of symptoms a small treatment effect of 0.30 was observed at 12-months (-4.9; 95% CI: -10.3, 0.5). When limited to those in the top 50% of depressive symptoms, treatment effects were even greater at three-months (Cohen’s *d*: 0.50) and 12-months (Cohen’s *d*: 0.74) months. Similarly, for PTSD treatment events were even greater at both timepoints and suggested a large effect at 12-months (3-Month Cohen’s *d*: 0.58; 12-Month Cohen’s *d*: 0.94).

**Table 4.**
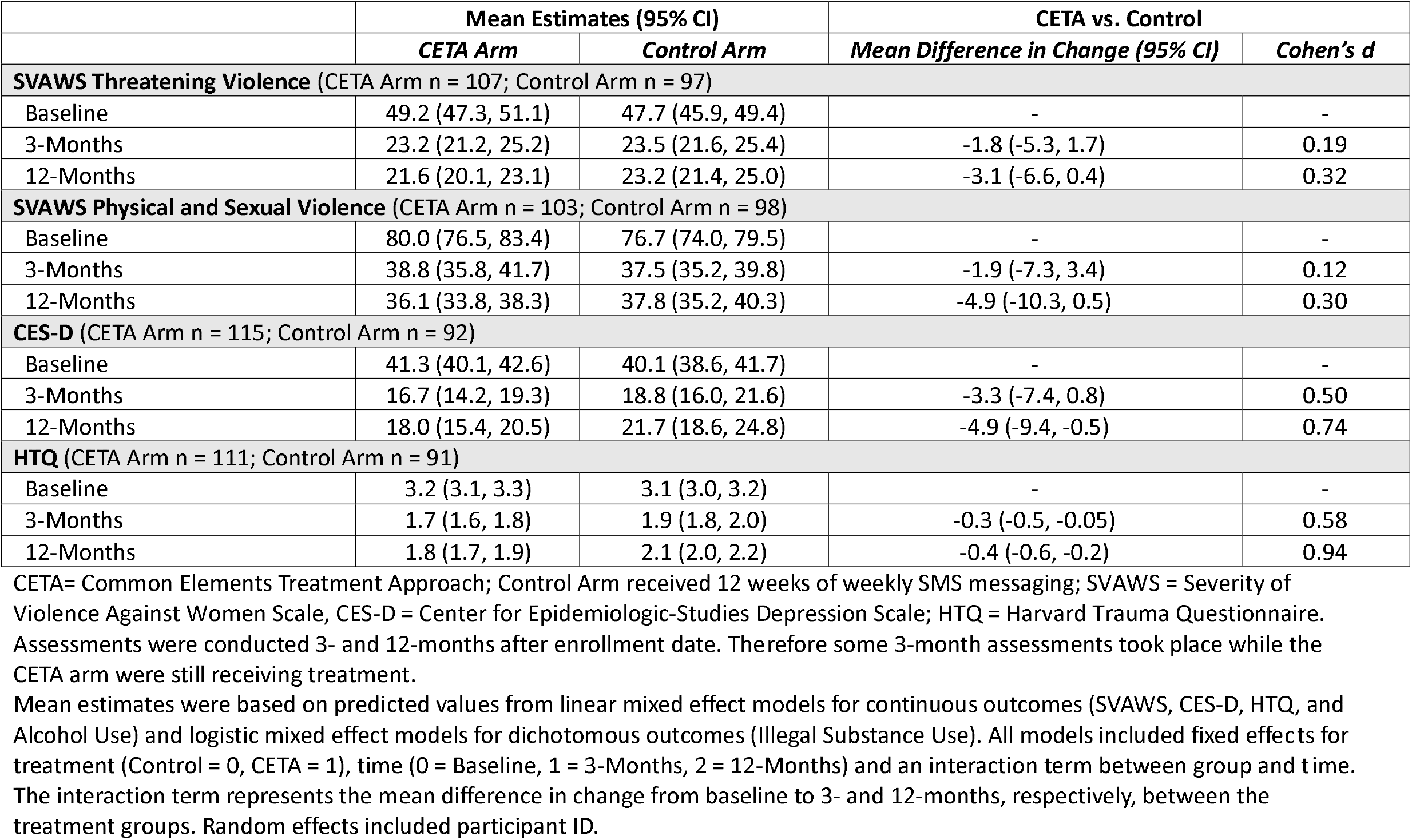
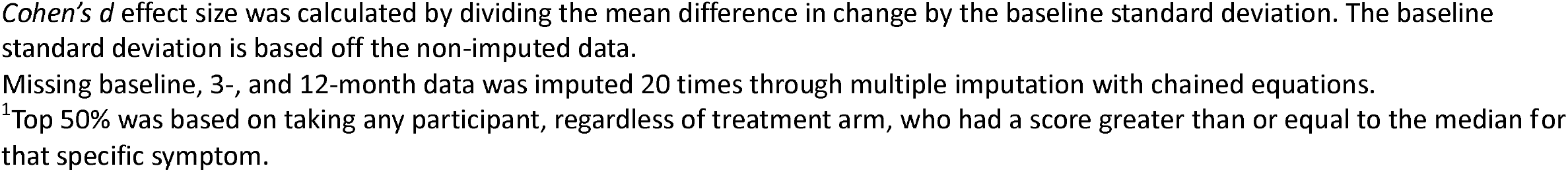
Subgroup Analysis: Estimated mean outcome differences and clinical effect sizes among participants in the top 50% of baseline symptoms in a randomized trial of the effect of CETA on mental health outcomes in Johannesburg South Africa among women with HIV on ART who have experienced violence and challenges with adherence ^1^

|  | Mean Estimates (95% CI) |  | CETA vs. Control |  |
| --- | --- | --- | --- | --- |
|  | CETA Arm | Control Arm | Mean Difference in Change (95% CI) | Cohen's d |
| **SVAWS Threatening Violence** (CETA Arm n = 107; Control Arm n = 97) |  |  |  |  |
| Baseline | 49.2 (47.3, 51.1) | 47.7 (45.9, 49.4) | - | - |
| 3-Months | 23.2 (21.2, 25.2) | 23.5 (21.6, 25.4) | -1.8 (-5.3, 1.7) | 0.19 |
| 12-Months | 21.6 (20.1, 23.1) | 23.2 (21.4, 25.0) | -3.1 (-6.6, 0.4) | 0.32 |
| **SVAWS Physical and Sexual Violence** (CETA Arm n = 103; Control Arm n = 98) |  |  |  |  |
| Baseline | 80.0 (76.5, 83.4) | 76.7 (74.0, 79.5) | - | - |
| 3-Months | 38.8 (35.8, 41.7) | 37.5 (35.2, 39.8) | -1.9 (-7.3, 3.4) | 0.12 |
| 12-Months | 36.1 (33.8, 38.3) | 37.8 (35.2, 40.3) | -4.9 (-10.3, 0.5) | 0.30 |
| **CES-D** (CETA Arm n = 115; Control Arm n = 92) |  |  |  |  |
| Baseline | 41.3 (40.1, 42.6) | 40.1 (38.6, 41.7) | - | - |
| 3-Months | 16.7 (14.2, 19.3) | 18.8 (16.0, 21.6) | -3.3 (-7.4, 0.8) | 0.50 |
| 12-Months | 18.0 (15.4, 20.5) | 21.7 (18.6, 24.8) | -4.9 (-9.4, -0.5) | 0.74 |
| **HTQ** (CETA Arm n = 111; Control Arm n = 91) |  |  |  |  |
| Baseline | 3.2 (3.1, 3.3) | 3.1 (3.0, 3.2) | - | - |
| 3-Months | 1.7 (1.6, 1.8) | 1.9 (1.8, 2.0) | -0.3 (-0.5, -0.05) | 0.58 |
| 12-Months | 1.8 (1.7, 1.9) | 2.1 (2.0, 2.2) | -0.4 (-0.6, -0.2) | 0.94 |

CETA= Common Elements Treatment Approach; Control Arm received 12 weeks of weekly SMS messaging; SVAWS = Severity of Violence Against Women Scale, CES-D = Center for Epidemiologic-Studies Depression Scale; HTQ = Harvard Trauma Questionnaire. Assessments were conducted 3- and 12-months after enrollment date. Therefore some 3-month assessments took place while the CETA arm were still receiving treatment.
Mean estimates were based on predicted values from linear mixed effect models for continuous outcomes (SVAWS, CES-D, HTQ, and Alcohol Use) and logistic mixed effect models for dichotomous outcomes (Illegal Substance Use). All models included fixed effects for treatment (Control = 0, CETA = 1), time (0 = Baseline, 1 = 3-Months, 2 = 12-Months) and an interaction term between group and time. The interaction term represents the mean difference in change from baseline to 3- and 12-months, respectively, between the treatment groups. Random effects included participant ID.
*Cohen's d* effect size was calculated by dividing the mean difference in change by the baseline standard deviation. The baseline standard deviation is based off the non-imputed data.
Missing baseline, 3-, and 12-month data was imputed 20 times through multiple imputation with chained equations.
<sup>1</sup>Top 50% was based on taking any participant, regardless of treatment arm, who had a score greater than or equal to the median for that specific symptom.

## DISCUSSION

This is the sixth clinical trial of CETA that has been conducted in a LMIC among a violence-affected population. In a prior trial of women experiencing ongoing IPV in Zambia by their current male partner, CETA was highly effective in reducing IPV and unhealthy alcohol use among the male partner with the trial stopped early by its DSMB due to effectiveness [15]. In Thailand, Iraq, Colombia, and Ukraine CETA was effective in reducing depression and PTSD symptoms compared to wait list control groups at post-treatment assessments [10–13]. In line with these prior trials, we similarly found strong effects of CETA for depression and PTSD here. The current study expands on those findings by having a stronger, active attention control condition and an extended follow-up timepoint at 12-months demonstrating long-term CETA impacts.

In contrast to the prior CETA trial on IPV, however, our intent-to-treat analysis found null or small effect sizes of CETA for threatened and physical/sexual IPV. There are several possible explanations for the smaller impact in this study. First, in this trial we included women who had experienced any violence in the past 12-months. In the Zambia trial, the inclusion criterion for IPV was past three-months and almost all of the women in the Zambia trial were experiencing ongoing IPV from their current male partner. For many women enrolled in the present study, they either did not have a current partner (22%) or were not cohabitating with their partner (55%), and the IPV they experienced had happened in the past and was not ongoing, which renders the SVAWS score less sensitive to change through an intervention like CETA. Enrolling participants with more recently experienced and/or ongoing IPV is recommended for future trials. Relatedly, in the Zambia trial all the women joined the study with their current male partners, who were the perpetrators of the IPV. In this trial, although we allowed male partners to be included, we only enrolled 15 male partners. A primary mechanism through which IPV was reduced in the Zambia trial was by a reduction in unhealthy alcohol use among male partners; in this study, CETA was largely only delivered to women, which precluded a key mechanism through which CETA was previously found to be effective. Taking these two trials together, the disparate findings suggest that engagement of men in IPV reduction interventions is important, as has been argued, but rarely implemented [26].

Similar to results from prior CETA trials, we found strong effect sizes for depression and PTSD. There were two novel CETA findings from this trial for these outcomes. First, we found that CETA treatment effects were larger among participants with the most severe symptoms. CETA is designed to address not only a range of mental health problems, but also a range of symptom severity [9]. This distinguishes the intervention from others that are designed to treat lower threshold mental health problems and often have stricter exclusion criteria (i.e., excluding individuals with the most severe problems) [27–29]. Second, this was the first time CETA treatment effects on depression and PTSD were evaluated at an extended follow-up—12 months post-baseline. Prior CETA trials and most global mental health trials have been limited by short follow-up periods. We found that CETA treatment effects were actually larger at 12 months than at three months for depression and PTSD. Reduced three-month effects may be attributed to a majority (54.8%) of the treatment cases either not completing CETA treatment prior to or completing CETA on the same day as the assessment timepoint. Our findings thus collectively suggest that CETA is effective for depression and PTSD among both milder and more severe levels and treatment effects are largely sustained over at least a year.

Our findings should be interpreted within the context of several limitations. First, findings for our substance use outcome should be considered exploratory in nature given the low prevalence of substance use at baseline as we were underpowered for this outcome. Second, this trial utilized self-reported assessment of all outcomes, and they are thus subject to measurement error, although the use of ACASI likely reduced the risk of social desirability. Third, much of the IPV reported in this trial was considered historical in nature – many of the women who enrolled in the study were no longer with the male partner who perpetrated the violence. Simultaneously, we did not require a male partner to enroll in CETA. Therefore, a combination of these factors limited our ability to detect clinical effects for the outcome of IPV.

## Supporting information

CONSORT

## Data Availability

Data generated by the study will be made available in de-identified format following protocol closure in a publicly available repository, to be identified in papers published from this study. Data obtained from the study sites (routinely generated medical record data) will not be owned by the authors and cannot be made available by them.

## CLINICAL IMPLICATIONS

Findings from this trial support the use of CETA to address issues related to trauma and depression among women living with HIV who reported experiences of IPV in the last 12-months. This is especially needed given the limited availability of appropriate and affordable mental health treatment options in South Africa’s HIV care settings specifically and in LMIC more broadly. Further research is needed to definitively evaluate CETA’s effectiveness for substance use. While treatment effects for IPV were small, the evidence from this trial and the prior trials of CETA suggest that the use of CETA is an effective approach to addressing IPV, however, it is important to engage and include male partners when possible. Furthermore, the women enrolled in our study were older and so the effectiveness of CETA may differ by age group where younger women may be more likely exposed to violence and more affected by such violence.

---

**Supplemental Figure 1.**
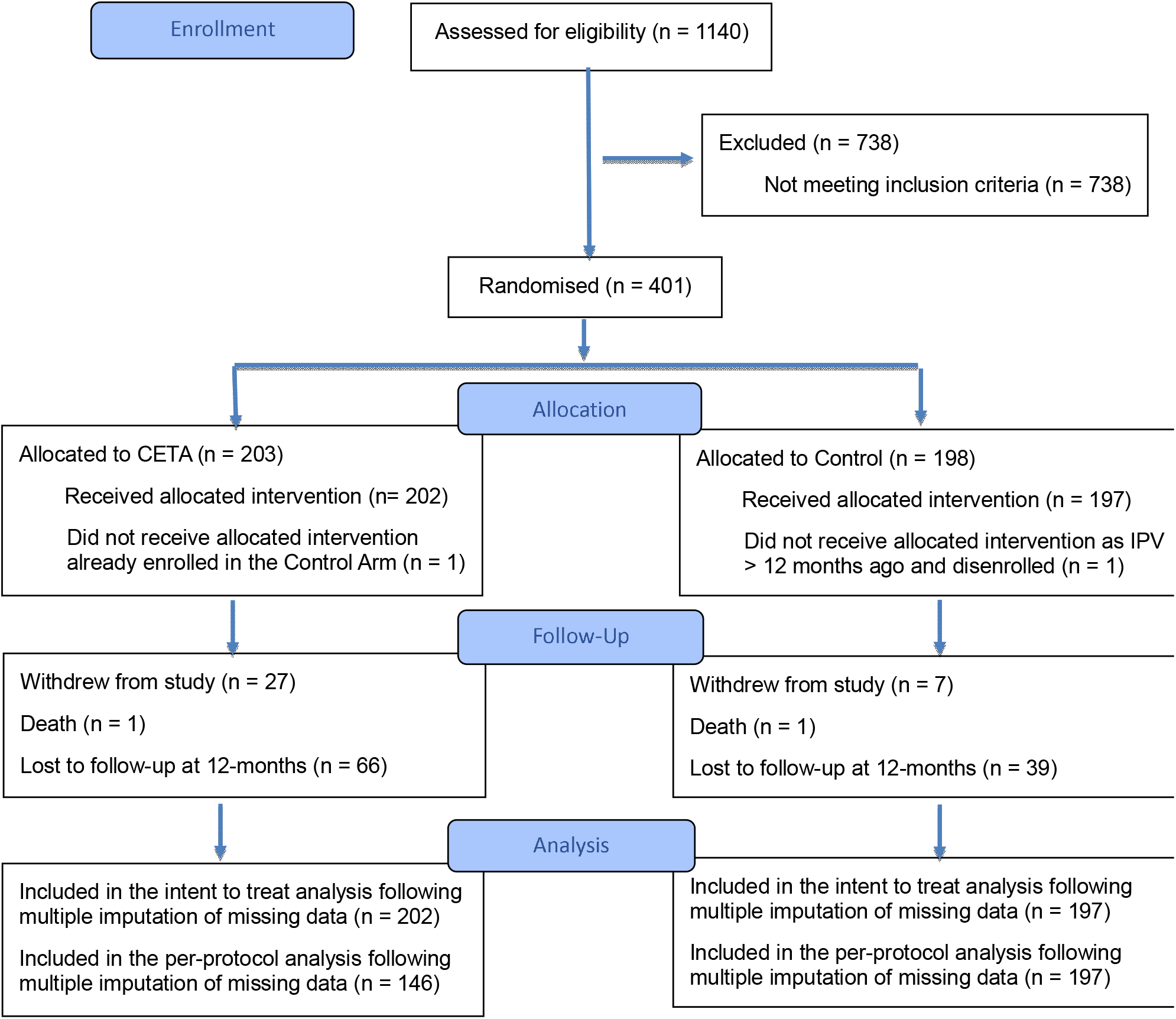
CONSORT Diagram Depicting the Flow of Participants Through the Phases of the CETA Trial

